# Effects of Different Intensity of Work Physical Activity on Sleep Disorder in Patients with Cholesterol

**DOI:** 10.1101/2023.10.26.23297614

**Authors:** Yiwen Cao, Rui Feng

## Abstract

**Background:** High cholesterol level is an important risk factor for cardiovascular disease. We believe that it is important to improve sleep disorders in patients with hyperlipidemia by exploring the effects of different intensities of physical activity on patients with hyperlipidemia.

**Objective:** The purpose of this study is to explore the relationship between sleep disorder and different intensity of work physical activity in patients with high cholesterol.

**Method and Materials:** This study used a randomly selected American population from the National Health and Nutrition Survey (NHANES) from 2017 to 2018 as the survey sample, consisting of 1515 individuals (770 males and 745 females, with an average age of 60.53 ± 14.232 years). The Categorical variable uses the Chi-squared test, and the measurement variable uses the rank sum test to calculate the test statistics, which is considered statistically significant (two-sided test) with P<0.05. After single factor analysis, we take physical activity at work as independent variable, take statistically significant variables in Demography (gender, race, marital status, income poverty ratio, physical activity, current health status) as covariates, and take sleep disorder as dependent variable to conduct binary logistic regression analysis.

**Results:** After excluding all the confounding factors, there was a significant correlation between work physical activity and sleep disorder (P<0.001), and the OR was 1.251 (95% Cl: 1.096-1.429).

**Conclusion:** Physical activity at work is a risk factor for sleep disorder in patients with high cholesterol, and the increased risk of sleep disorder caused by intense physical activity at work is particularly obvious.

## 1 Research background

### 1.1 The risk of high cholesterol levels

According to the global high cholesterol population change and driving force report published by the British journal ⟪Nature⟫, about 1000 researchers around the world analyzed 1127 research data and concluded that the population of Hypercholesterolemia in low - and middle-income countries continued to increase from 1980 to 2018. At the same time, changes in diet, physical activity and treatment methods are important causes of Hypercholesterolemia. High cholesterol level is an important risk factor for cardiovascular disease, especially low-density lipoprotein cholesterol will increase the damage to arteries. Therefore, in 2017, low-density lipoprotein cholesterol was considered to be one of the factors affecting 3.9 million deaths worldwide. (1)Due to pathological and physiological changes, patients with high cholesterol will suffer from malignant development of cholesterol levels, cardio cerebral Vascular disease, joint inflammation, and eye diseases. These diseases and symptoms will increase patients’ suffering from sleep disorder.

### 1.2 The relationship between high cholesterol level and sleep disorder

In 2008, Miedema HM et al. conducted a study on serum cholesterol and sleep duration in elderly people and found that longer sleep duration was associated with higher total cholesterol levels.(2) In 2017, Michael K Lemke et al. conducted a study on “the cholesterol level and sleep of long-distance truck drivers in the United States”, which showed that long-distance truck drivers have a high cholesterol risk, and sleep quality is one of the factors inducing cholesterol risk, that is, sleep quality is closely related to High-density lipoprotein, low-density lipoprotein and total cholesterol. (3) The research results of Niloufar Rasaei et al. on the cholesterol/Saturated fat acid index CSI and sleep quality of overweight and obese women in 2023 show that people with higher CSI units are more likely to have sleep problems. (4) We can see that high cholesterol levels have a certain impact on the sleep of different people, causing insomnia, sleep apnea syndrome and other diseases, while sleep disorder will induce the risk of high cholesterol, causing Atherosclerosis and other diseases, seriously affecting people’s health. Therefore, we chose the special population of high cholesterol patients as the research object.

### 1.3 The relationship between physical activity and high cholesterol

Hypercholesterolemia is an important health problem worldwide. Many studies have confirmed that active physical activity can improve the level of HDL-C, which is conducive to the reverse operation of cholesterol and regulates the level of human cholesterol and blood lipids. Finally, change the problem of increased incidence rate and mortality caused by high cholesterol levels. (5) However, due to the unique physiological and pathological characteristics of high cholesterol patients, not all types of physical activities are suitable for high cholesterol patients, and their physical activities should be distinguished from those of the general population. (6) It is necessary to conduct targeted research on the exercise patterns, intensity, and frequency of high cholesterol individuals related to physical activity arrangements. Therefore, the pathological and physiological effects on patients with high cholesterol still deserve further attention and research. This study aims to explore whether non Exercise prescription intervention can bring positive effects to high cholesterol patients by studying the intensity of patients’ physical activities at work.

### 1.4 The relationship between physical activity and sleep disorder

Sleep is a cyclical activity in human life. With the impact of life pressure and excessive use of electronic devices, people of all ages are troubled by sleep disorder and insomnia. Severe sleep disorder is a risk factor leading to mental illness, cardiovascular disease and other diseases. Numerous studies have shown that physical activity has a significant impact on sleep quality. The research results encourage people to increase their physical activity appropriately. (7) At the same time, according to the “Physical Activity Guidelines for the Chinese Population (2021)”, people with chronic diseases and the elderly should engage in regular physical activities based on their own physical conditions, reduce sedentary time, improve bad living habits, and improve sleep quality. (8)Therefore, in order to improve sleep disorder of patients with high cholesterol by improving physical activity, the first task is to study whether physical activity is a protective factor for their sleep disorder. In this way, we can avoid the secondary damage to the body of patients with high cholesterol caused by excessive fatigue after physical activity, and formulate Exercise prescription suitable for patients with high cholesterol, so as to reduce patients’ troubles.

## 2 Objects and Methods

### 2.1 Research subjects

This study focuses on a random sampling survey of 9254 people conducted by the National Health and Nutrition Survey (NHANES) in the United States from 2017 to 2018 to understand the health and nutritional status of the entire population. (9) According to the research direction of “physical activity and sleep disorder of patients with high cholesterol”, a total of 1515 patients with high cholesterol were selected as research objects based on the data of Demographics, physical activity, cholesterol level and sleep disorder. The survey plan and secondary analysis of the data were approved by the Ethics Review Committee of the National Center for Health Statistics in the United States, providing written consent notices for all adult participants.(10) Inclusion criteria: ➀ Adult residents aged 20 and above in the United States; ➁ Able to independently fill out questionnaires and consciously answer interviewees’ questions; ➂ Agree to cooperate in completing the entire investigation process. Exclusion criteria: ➀ Age does not meet the criteria; ➁ Individuals with unclear awareness or inability to engage in normal conversations with interviewees; ➂ Refusing to cooperate in completing the entire investigation process. (See Figure 1)

**Figure 1.**
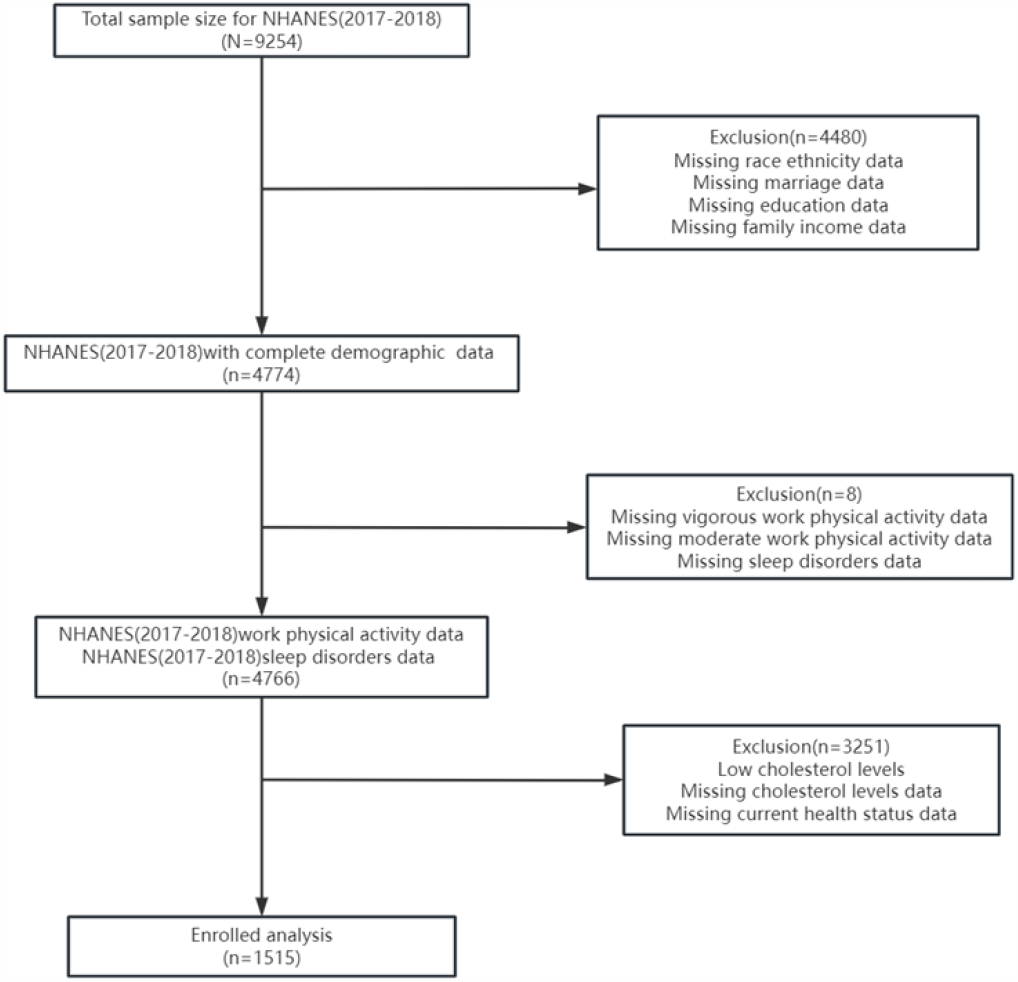
Data Screening Flowchart

### 2.2 Evaluation of Demography Variables

This study belongs to a cross-sectional survey, and 9254 people were randomly sampled across the United States to participate in the survey. We selected gender, age, race, education level, marital status, income poverty ratio, and current health status as demographic variables. Among these variables, age is divided into 20-39 years old, 40-59 years old, and over 60 years old. Race is divided into Mexican Americans, other Hispanic, non Hispanic white, non Hispanic black, non Hispanic Asian, and other races. Education level is divided into high school education below, high school graduation, and high school education above. Marital status is divided into cohabitation, married living alone, and never married. The income poverty ratio is the ratio of household income to the poverty criteria of the survey year. In this study, the poverty ratio is divided into two types: poverty (<1.3) and middle-income (≥ 1.3). (11,12)

### 2.3 Assessment of physical activity, high cholesterol and sleep disorder

The NHANES physical activity questionnaire for 2017-2018 was designed based on the Global Physical Activity Questionnaire (GPAQ). In the questionnaire, we selected two items: intense work activity (PAQ605) and moderate work activity (PAQ620) as the basis for evaluating the intensity of work physical activity among respondents. The respondents will provide corresponding answers based on whether their paid or unpaid work (work, household chores, or other) within a week has caused a significant or slight increase in breathing or heart rate, and the interviewee will record and evaluate the intensity of physical activity. According to the survey results, in paid or unpaid work, we classify physical activity at work into three categories: 1=intense physical activity; 2=moderate intensity of physical activity, but not intense physical activity; 3=No physical activity.

The blood pressure and cholesterol dataset of the 2017-2018 NHANES questionnaire includes ten questions about blood pressure and cholesterol, among which there are five questions about cholesterol levels. We chose “diagnosed with high cholesterol by medical institutions” (BPQ080) in the questionnaire as the criterion for determining whether cholesterol levels are high or low.

The 2017-2018 NHANES questionnaire sleep disorder data set includes ten questions about sleep habits and disorders on weekdays and weekends. We chose “whether to tell doctors about sleep disorder” (SLQ050) in the questionnaire as the basis for this study to judge whether there is sleep disorder.

### 2.4 Quality Control

To ensure the quality of data collection, all interviewers need to strictly follow the Interviewer Procedure Manual when collecting data. The interviewer uses a computer-assisted personal interview (CAPI) system to ask questions, and the CAPI system is programmed with built-in consistency checks to reduce data input errors; CAPI uses the ‘online help screen’ to help interviewers define key terms used in survey questionnaires. In addition, to ensure the accuracy and completeness of data, NHANES field office staff will conduct rigorous reviews.

### 2.5 Statistical Analysis

This study used Microsoft Excel 2010 to sort out and summarize the data of Demography, cholesterol level, physical activity and sleep disorder collected by NHANES from 2017 to 2018, including eliminating invalid data and missing data (rejection and unknown) items, and classifying and assigning values to the data. According to the purpose of the study, statistical testing was conducted on the data using the Statistical Package for Social Sciences (SPSS) version 26.0 analysis software.

We conducted a significant test on the difference of covariates of “sleep disorder”, used Chi-squared test for Categorical variable, and used rank sum test for continuous variables to select statistically significant factors. In the univariate analysis, the variables with statistical significance were included in the stepwise binary logistic regression analysis. The binary logistic regression model was used to analyze the relationship between “work physical activity and sleep disorder in patients with high cholesterol”. The P value less than 0.05 was considered statistically significant (bilateral test). Select and exclude confounding variables using A-entry=0.05 and a-exit=0.10.

Taking physical activity at work as independent variable and sleep disorder as dependent variable, in single factor analysis, gender, race, marital status, income poverty ratio and current health status were statistically significant (P<0.05); Age (P=0.527) and education level (P=0.512) were not statistically significant. When analyzing the relationship between work physical activity and sleep disorder, in order to exclude the influence of mixed factors, we established the following models for single factor and multi factor binary logistic regression analysis. Model I: only adjusted for the independent variable of physical activity; Model II: adjusted by adding Demography variables according to the independent variables in model I.

## 3 Data analysis

### 3.1 Demographics characteristics

According to the research purpose and data validity, this study screened and sorted out the data collected by the National Health and Nutrition Survey Center (NHANES) in 2017-2018, and finally obtained the Demography and work physical activity related data of 1515 patients with high cholesterol. The participants in the survey included 770 males and 745 females, with an average age of 60.53 ± 14.232 years. There were 572 participants with sleep disorder and 943 participants without sleep disorder. Between the sleep disorder group and the sleep disorder free group, in the univariate analysis, we found that the patients with high cholesterol had statistical significance in gender (P=0.003), race (P=0.000), marital status (P=0.008), income poverty ratio (P=0.009), current health status (P=0.000) and physical activity at work (P=0.011). Age (P=0.206) and education level (P=0.512) were not statistically significant. (See Table 1)

**TABLE 1.**
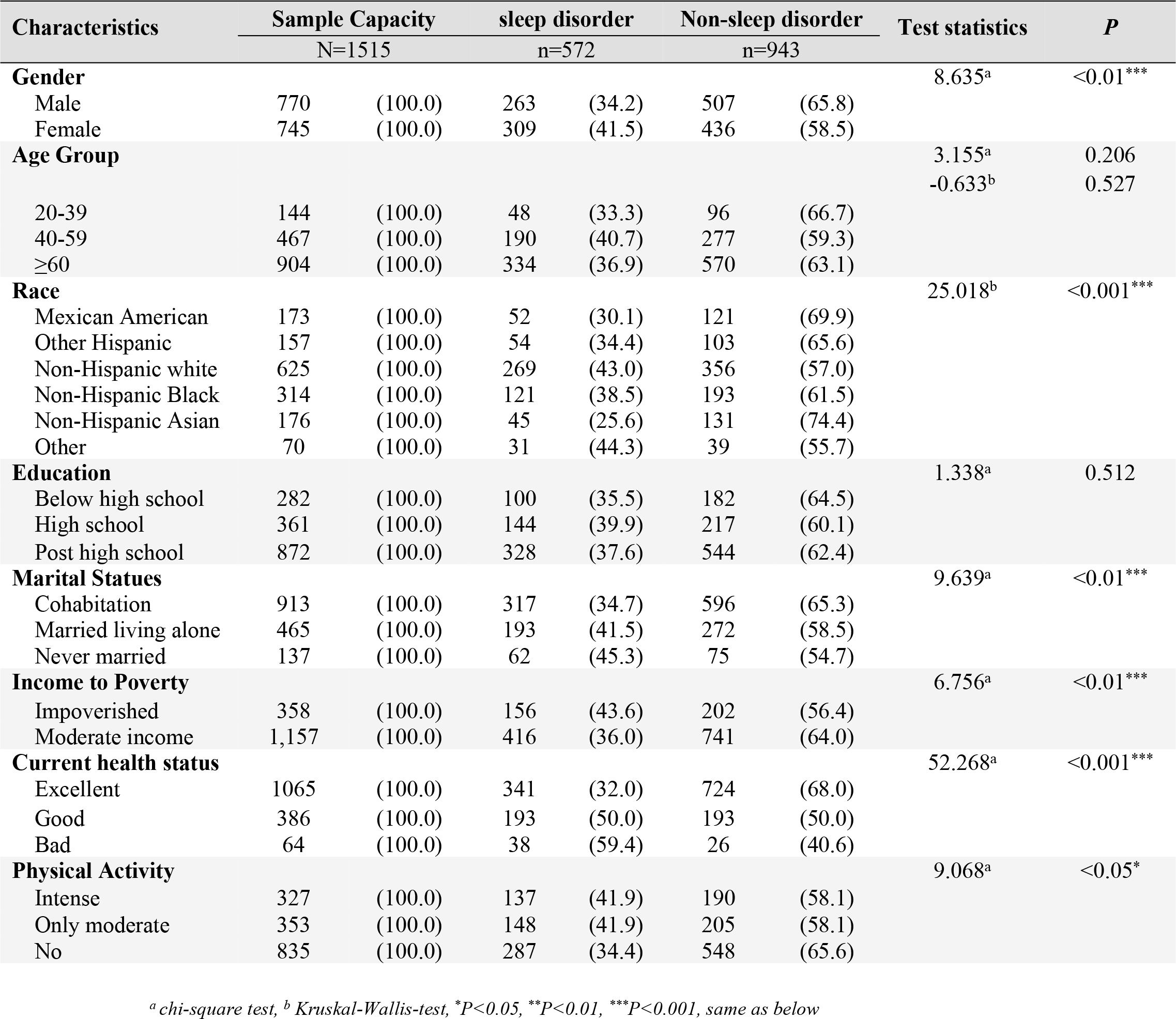
Demographic characteristics of patients over age with high cholesterol.

| Characteristics | Sample Capacity |  | sleep disorder |  | Non-sleep disorder |  | Test statistics | P |
| --- | --- | --- | --- | --- | --- | --- | --- | --- |
|  | N=1515 |  | n=572 |  | n=943 |  |  |  |
| **Gender** |  |  |  |  |  |  | 8.635<sup>a</sup> | <0.01*** |
| Male | 770 | (100.0) | 263 | (34.2) | 507 | (65.8) |  |  |
| Female | 745 | (100.0) | 309 | (41.5) | 436 | (58.5) |  |  |
| **Age Group** |  |  |  |  |  |  | 3.155<sup>a</sup> | 0.206 |
|  |  |  |  |  |  |  | -0.633<sup>b</sup> | 0.527 |
| 20-39 | 144 | (100.0) | 48 | (33.3) | 96 | (66.7) |  |  |
| 40-59 | 467 | (100.0) | 190 | (40.7) | 277 | (59.3) |  |  |
| ≥60 | 904 | (100.0) | 334 | (36.9) | 570 | (63.1) |  |  |
| **Race** |  |  |  |  |  |  | 25.018<sup>b</sup> | <0.001*** |
| Mexican American | 173 | (100.0) | 52 | (30.1) | 121 | (69.9) |  |  |
| Other Hispanic | 157 | (100.0) | 54 | (34.4) | 103 | (65.6) |  |  |
| Non-Hispanic white | 625 | (100.0) | 269 | (43.0) | 356 | (57.0) |  |  |
| Non-Hispanic Black | 314 | (100.0) | 121 | (38.5) | 193 | (61.5) |  |  |
| Non-Hispanic Asian | 176 | (100.0) | 45 | (25.6) | 131 | (74.4) |  |  |
| Other | 70 | (100.0) | 31 | (44.3) | 39 | (55.7) |  |  |
| **Education** |  |  |  |  |  |  | 1.338<sup>a</sup> | 0.512 |
| Below high school | 282 | (100.0) | 100 | (35.5) | 182 | (64.5) |  |  |
| High school | 361 | (100.0) | 144 | (39.9) | 217 | (60.1) |  |  |
| Post high school | 872 | (100.0) | 328 | (37.6) | 544 | (62.4) |  |  |
| **Marital Statues** |  |  |  |  |  |  | 9.639<sup>a</sup> | <0.01*** |
| Cohabitation | 913 | (100.0) | 317 | (34.7) | 596 | (65.3) |  |  |
| Married living alone | 465 | (100.0) | 193 | (41.5) | 272 | (58.5) |  |  |
| Never married | 137 | (100.0) | 62 | (45.3) | 75 | (54.7) |  |  |
| **Income to Poverty** |  |  |  |  |  |  | 6.756<sup>a</sup> | <0.01*** |
| Impoverished | 358 | (100.0) | 156 | (43.6) | 202 | (56.4) |  |  |
| Moderate income | 1,157 | (100.0) | 416 | (36.0) | 741 | (64.0) |  |  |
| **Current health status** |  |  |  |  |  |  | 52.268<sup>a</sup> | <0.001*** |
| Excellent | 1065 | (100.0) | 341 | (32.0) | 724 | (68.0) |  |  |
| Good | 386 | (100.0) | 193 | (50.0) | 193 | (50.0) |  |  |
| Bad | 64 | (100.0) | 38 | (59.4) | 26 | (40.6) |  |  |
| **Physical Activity** |  |  |  |  |  |  | 9.068<sup>a</sup> | <0.05* |
| Intense | 327 | (100.0) | 137 | (41.9) | 190 | (58.1) |  |  |
| Only moderate | 353 | (100.0) | 148 | (41.9) | 205 | (58.1) |  |  |
| No | 835 | (100.0) | 287 | (34.4) | 548 | (65.6) |  |  |

<sup>a</sup> chi-square test, <sup>b</sup> Kruskal-Wallis-test, \* $P<0.05$ , \*\* $P<0.01$ , \*\*\* $P<0.001$ , same as below

### 3.2 Logistic regression analysis of different intensity work physical activity and sleep disorder in patients with high cholesterol

In Logistic regression analysis, model I (without excluding any confounding variables) showed that the odds ratio (OR) associated with physical activity and sleep disorder at work was 1.195 (95% CI: 1.052-1.357); Model II (excluding Demography variables) shows that the OR value is 1.251 (95% CI: 1.096-1.429). The research results show that after adjusting all the confounding factors, the Logistic regression is a Nonlinear regression, and the role of the independent variable “physical activity” should be determined by the OR value. In this study, the regression coefficient of “physical activity” is a positive value (b=0.224), which can be understood as the increase of physical activity intensity during work, and the risk of sleep disorder will increase. Physical activity at work is a risk factor for sleep disorder in patients with high cholesterol, and the correlation between physical activity at work and high risk of sleep disorder increases. The risk of sleep disorder in patients with high cholesterol who have physical activity at work is 1.251 times that of those who do not have physical activity at work (P<0.01). (Table 2)

**Table 2.**
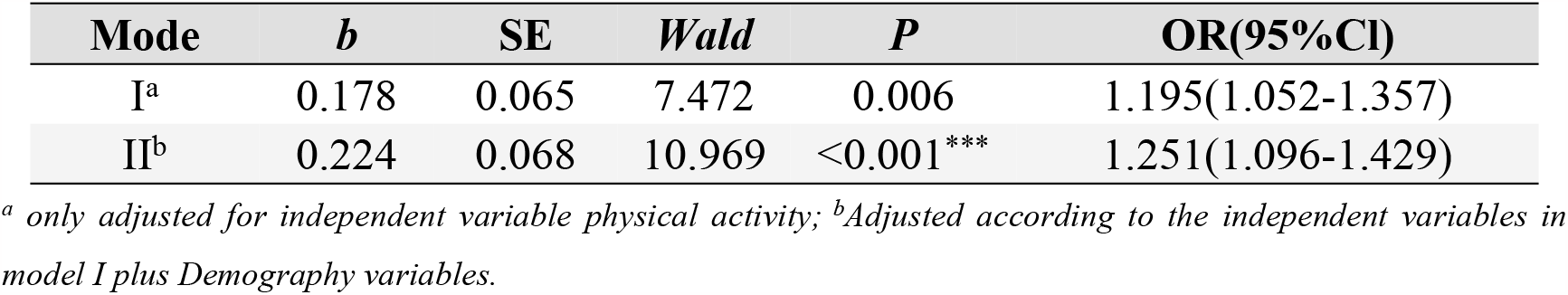
Correlation between physical activity at work and sleep disorder.

In this study, physical activity at work is divided into three levels, so we use multiple logistic regression analysis to further analyze the impact of different intensity physical work activities at work on sleep disorder. The research results show that for people with high cholesterol and sleep disorder, “intense physical activity” is 1.444 times of “only moderate physical activity” in work (p<0.01), and “only moderate physical activity” is 1.375 times of “no physical activity” in work (p<0.05). Therefore, with the increase of physical activity intensity at work, the risk of sleep disorder in patients with high cholesterol increases, and the risk of sleep disorder caused by intense physical activity is the highest. (See Table 3)

**Table 3.** Effects of physical activity on sleep disorder in different intensity of work.

| Physical activity | *b* | SE | Wald | *P* | OR(95%CI) |
| --- | --- | --- | --- | --- | --- |
| Intense physical activity | 0.368 | 0.141 | 6.772 | 0.009 | 1.444 (1.095-1.905) |
| Only moderate intensity physical activity | 0.318 | 0.136 | 5.465 | 0.019 | 1.375 (1.053-1.796) |
| No physical activity* | 0<sup>b</sup> | . | . | . | . |

\* set as a reference.

## 4 Discussion

Through the logistic regression analysis of the data of NHANES from 2017 to 2018, we found that among the patients with high cholesterol aged over 20, physical activity at work was associated with sleep disorder. At the same time, by comparing intense physical activity with moderate physical activity at work, we found that with the increase of the intensity of physical activity at work, the risk of sleep disorder in patients with high cholesterol will increase accordingly, that is, high intensity physical activity at work is a risk factor for sleep disorder in patients with high cholesterol.

### 4.1 Work physical activity is a risk factor for sleep disorder in patients with high cholesterol

Sleep is an important process of body repair and energy recovery in the human body. In many studies on the relationship between physical activity and sleep, it has been concluded that physical activity is beneficial for improving sleep quality. And people with various clinical diseases have always been the focus of sleep disorder research. For example, whether daily physical activities of people with coronary heart disease, hypertension, high cholesterol, etc. will affect the sleep quality of clinical patients. Recent studies have shown that for patients with high cholesterol, a good sleep state has a positive effect on controlling the deterioration and development of their condition. As Aho V et al. confirmed in their study, sleep deprivation reduces the expression of genes encoding cholesterol transporters and can increase the expression of inflammatory response pathways. At the same time, the research report also pointed out that the High-density lipoprotein of sleep deprived subjects was low, while in the experimental sleep restriction study, the circulating low-density lipoprotein was reduced, and prolonged sleep disorder would change inflammation and cholesterol levels in terms of gene expression and serum lipoprotein levels. (13) The research of Sol Moch ó n-Benguigui et al. shows that shortening sedentary time and increasing physical activity and fitness time is an important way for middle-aged people to prevent or treat potential diseases, which is conducive to promoting the development of physical and mental health of patients and reducing sleep disorder. (14) Among many forms of lifestyle change, increasing physical activity is widely considered as an important way to reduce blood pressure and blood cholesterol, which can reduce the risk of physical diseases caused by elevated blood pressure and blood cholesterol, including the risk of “sleep disorder”. (15) Therefore, changes in sleep and physical activity are closely related to the health status of adults, and previous studies have suggested a bidirectional correlation. (16)

Although physical activity has been proved to have a positive impact on the physical quality of patients with Dyslipidemia, and can effectively reduce cholesterol levels (17), perhaps due to the impact of gender, age, race, sleep habits and other differences, there is less research on the correlation between physical activity at work and sleep disorder in patients with high cholesterol. And there is a certain degree of difference between the existing research conclusions and our research. Our research data shows that the increase of physical activity at work in patients with high cholesterol is positively related to the increase of the risk of sleep disorder, which means that the risk of sleep disorder will also increase if they participate in more vigorous physical activities at work.

For the results of data analysis of this study, “physical activity at work is a risk factor for sleep disorder in patients with high cholesterol, and the greater the intensity of physical activity, the greater the risk of sleep disorder”. We may explain it in three aspects: ➀ The physical activity patterns in work are not suitable for high cholesterol patients. For example, high-intensity continuous handling of heavy objects at work leads to a significant increase in heart rate or respiratory rate, resulting in an increase in oxygen consumption. Patients with high cholesterol may have plaques in their blood vessels due to pathological reasons, which can block blood flow and lead to insufficient blood oxygen supply; In patients with more serious symptoms, Atherosclerosis is triggered, leading to blood flow blockage. Excessive physical activity not only has no effect on the improvement of sleep disorder, but also directly endangers life due to cerebral ischemia in serious cases. ➁ The best exercise way to improve sleep disorder of patients with high cholesterol is high-intensity aerobic exercise and medium intensity resistance training. (18,19) The increase in calorie expenditure related to aerobic exercise has a positive impact on lowering cholesterol levels. It is recommended to start with moderate intensity aerobic exercise and target high intensity aerobic exercise to improve high cholesterol levels while also improving sleep quality. (20) At the same time, balancing resistance training can demonstrate more benefits for patients’ physiological and psychological systems. (21)➂ The sleep quality of patients with high cholesterol is not only affected by physical activity, but also related to factors such as sleeping habits, eating habits and living environment. The research on the correlation between the above factors and sleep disorder of patients with high cholesterol is also worth further confirmation.

Related studies have shown that a combination of moderate intensity resistance training and aerobic training has significant effects on improving body composition, lipid metabolism, blood sugar control, blood pressure, cardiovascular health, and muscle strength in high cholesterol individuals. (22)Therefore, if patients with high cholesterol want to improve their sleep disorder through physical activity, they need to comprehensively evaluate their physiological and biochemical indicators and formulate corresponding Exercise prescription. How to balance physical activities at work and daily fitness time for patients with high cholesterol, and how to formulate effective Exercise prescription for patients with high cholesterol to help them improve sleep disorder, are also another important direction proposed through this study. Based on this study, we suggest that patients with high cholesterol maintain a low intensity of physical activity during work and activities as much as possible to avoid causing harm to the body due to factors such as insufficient blood supply to the body; When engaging in non work physical activities, in order to avoid excessive exercise, physical function exercises should be carried out according to the actual physical condition, thereby promoting the improvement of sleep quality. If there is no condition to get professional guidance from doctors or coaches, patients with high cholesterol still need to maintain a low intensity in work physical activities. When doing non work physical activities, we recommend participating in activities with low intensity and sufficient oxygen supply, such as Taijiquan and yoga, within the safe range of exercise to improve sleep disorder.

### 4.2 Limitations of This Study

Regarding the limitations of this study, we can summarize them as follows: Firstly, the data we used were relevant survey data from US residents, and there is some controversy over whether the research findings can be extended to other countries and regions around the world. Secondly, regarding the strength of physical activity at work, our classification is relatively single, and we only use whether there is a significant increase in breathing or heart rate during work as a criterion to evaluate the intensity of physical activity. Third, the factors that lead to sleep disorder are diverse. This study only studied physical activity at work, and failed to exclude environmental factors, genetic factors and other factors. Fourthly, this study extracted data from 2017 to 2018, with a small sample size and a short time period. Future research designs plan to include longer period data with a larger sample size.

## 5 Conclusions

Our research results show that the increased intensity of physical activity at work is related to the increased risk of high cholesterol sleep disorder, which provides a scientific basis for people with Hypercholesterolemia to engage in physical activity and receive active exercise intervention.

## Data Availability

no

## Abbreviation

NHANES: National Health and Nutritional Examination Survey
OR: Odds Ratio
P: P-Value
U.S: The United States of America
CAPI: Computer-Assisted Personal Interview
GPAQ: Global Physical Activity Questionnaire
SPSS: Statistical Package for Social Sciences

## Declarations

### Ethics approval and consent to participate

All procedures performed in the study were in accordance with the Declaration of Helsinki. The study protocols for NHANES were approved by the National Center for Health Statistics (NCHS) Research Ethics Review Board (Protocol#2017–1). All adult participants provided written notification of consent before participating in the study.

### Consent for publication

Not applicable.

## Availability of data and materials

The datasets generated and/or analysed during the current study are available in the [NHANES] repository, [NHANES Questionnaires, Datasets, and Related Documentation (cdc.gov)]. Raw data supporting the obtained results are available at the corresponding author.

## Author Contributions

YC and RF conceived and designed the study. YC organized the database, performed the statistical analysis and wrote the manuscript. Linguist RF confirmed the accuracy of the written language. YC and RF revised the manuscript. All authors edited, revised, and certified the final version of this manuscript.

## Funding

This study were supported by the 2019 Sports Research Project of Henan Province Sports Bureau (Grants No. 2019052) and the 2020 General Project of Henan Province Education Science “13th Five-Year Plan” (Grants No. 2020YB0014).

## Conflict of Interest

The authors declare that the research was conducted in the absence of any commercial or financial relationships that could be construed as a potential conflict of interest.

## Acknowledgments

We would like to thank all the staff and participants of the National Health and Nutrition Examination Survey 201-2018 cycles for their valuable contributions. Any interpretation or conclusion related to this manuscript does not represent the views of the NHANES. We would also like to thank the editors and reviewers for their valuable and constructive comments to help us improve the manuscript. And we especially thank Dr. Feng for his constructive comments.

## Notes

### Clinical Trial

no

### Funding Statement

The author(s) received no specific funding for this work.

